# Bio-Inspired Attentive Segmentation of Retinal OCT imaging

**DOI:** 10.1101/2020.08.13.20174250

**Authors:** Georgios Lazaridis, Moucheng Xu, Saman Sadeghi Afgeh, Giovanni Montesano, David Garway-Heath

## Abstract

Albeit optical coherence imaging (OCT) is widely used to assess ophthalmic pathologies, localization of intra-retinal boundaries suffers from erroneous segmentations due to image artifacts or topological abnormalities. Although deep learning-based methods have been effectively applied in OCT imaging, accurate automated layer segmentation remains a challenging task, with the flexibility and precision of most methods being highly constrained. In this paper, we propose a novel method to segment all retinal layers, tailored to the bio-topological OCT geometry. In addition to traditional learning of shift-invariant features, our method learns in selected pixels horizontally and vertically, exploiting the orientation of the extracted features. In this way, the most discriminative retinal features are generated in a robust manner, while long-range pixel dependencies across spatial locations are efficiently captured. To validate the effectiveness and generalisation of our method, we implement three sets of networks based on different backbone models. Results on three independent studies show that our methodology consistently produces more accurate segmentations than state-of-the-art networks, and shows better precision and agreement with ground truth. Thus, our method not only improves segmentation, but also enhances the statistical power of clinical trials with layer thickness change outcomes.

## 1 Introduction

Optical coherence tomography (OCT) is a non-invasive imaging modality that provides high-resolution scans of the structures of the human retina [1]. The retina is organized into layers and, clinically, OCT is used as a surrogate measure to evaluate retinal cell loss by measuring layer thicknesses around the optic nerve head. Thus, OCT enables us to extract this depth information from retinal layers, which is known to change with certain ophthalmic pathologies, i.e. retinal nerve fibre layer (RNFL) thickness for glaucoma, and is also associated with neurodegenerative and vascular disorders [2]. Therefore, accurate and precise segmentation of retinal layers is necessary to assess morphological retinal changes in order to quantify presence or progression of pathologies.

OCT layer segmentation has produced a veritable soup of methodologies trying to address this challenging task. Classical approaches attempt to formulate the problem as a topologically correct graph or as an optimization problem based on a set of predefined or heuristic rules [3,4,5,6,7]. While these methods achieve remarkable results, their segmentation efficiency is limited in the presence of noise and artifacts, and results are highly sensitive to the choice of initial parameters. Moreover, topological continuity and smoothness in the obtained surfaces is not always guaranteed. Meanwhile, various methods using convolutional neural networks (CNNs) have been proposed to segment retinal OCT images [8,9,10,11,12,13]. For example, in [8], CNNs have been used to segment retinal layers by modeling the position distribution of the surfaces and by using a soft-argmax method to infer the final positions. In [9], layer segmentation is achieved by extracting the boundaries from probability maps and using a shortest path algorithm to obtain the final surfaces. The authors in [10] employ a modification of the encoder-decoder paradigm to produce dense predictions for every vertical column in each slice of the OCT volume, trying to maintain spatial correlation, whereas in [13], the authors use a U-Net [14] with residual blocks and diluted convolutions to achieve retinal layer segmentation. In [11], the authors propose to segment layers by classifying each pixel into layer or background based on an hierarchy of contextual features. In [12], segmentation is achieved by uniformly dividing the image into strips and then decomposing them into a sequence of connected regions.

These works may, however, present important limitations for OCT layer segmentation. Firstly, the previous approaches have inconsistent prediction boundaries which may not have spatial continuity. Secondly, signal and noise properties in OCT images occur at different spatial scales and, therefore, these methods might not be able to capture all the necessary information needed for segmentation. Finally, the specific geometry of OCT images is not fully exploited, thus reducing the probability for accurate and topologically sound segmentations. Therefore, principled schemes accounting for boundary morphology and signal topology must be developed, in order to preserve anatomical information and allow for spatial coherency.

This paper presents a novel end-to-end trainable method to improve retinal layer segmentation. Our methodology uses efficient high-order attention descriptors leveraging on the specific anatomical OCT geometry to extract robust quantifications of all retinal layers. Our model increases feature correlation and expression learning, exploiting the horizontally-layered retinal structure and the biological knowledge that retinal surfaces can be modeled as partitioned layers along the vertical dimension. We showcase the diagnostic precision and agreement of our method with ground truth RNFL (commonly assessed layer) segmentations from two independent studies [1,15]. Finally, we demonstrate the superiority of our method in segmenting all retinal layers using the Duke dataset [16].

## 2 Methods

### 2.1 Bio-Inspired Attentive Segmentation

OCT images have a very specific geometry, where layers and retinal boundaries are oriented along the horizontal and vertical directions. For this reason, we conjecture that segmentation tasks on these type of images can benefit from exploiting the orientation of the extracted features. Also, ignoring these structural priors aggravates the issue of topological inconsistencies and incorrect pixel classifications near the layer boundaries that OCT segmentation models often suffer from [11,17,13].

Instead of mathematically formulating prior anatomical knowledge or information around layer edges, we propose a method that implements these topological priors by constraining the orientation of the feature extraction layers -that is to say, by constraining the receptive field of the convolutional layers to focus separately on each direction. The features are, then, combined into an attention mask used to enhance the supervision signal for the segmentation task. Thus, our model is better able to extract features that are primarily oriented in the horizontal and vertical direction.

### 2.2 Low-Rank Oriented Attention (LROA)

Given an input tensor ***X*** ∈ *R^S×S×C^* and a parametrized bilinear weight matrix ***W*** ∈ ℝ*^N^ ^×M^*, the output is given by:

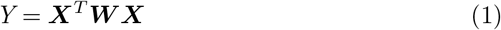

where *Y* ∈ ℝ*^N^* ^×^ *^M^*. Although pair-wise dependencies of discrete pixels (Eq. 1) are typically modelled as a non-local mean operation [18], the resulting computational cost is very high due to high-rank matrix product multiplications. To enable cost-efficient computing, we model spatial correlations using a low-rank approximation based on the Hadamard product [19]:

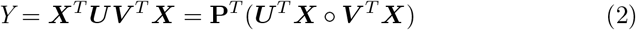

where o denotes the Hadamard product. Bias terms are omitted for simplicity. In the original formulation [19], ***U***, ***P***, ***V*** are linear projections. To incorporate prior anatomical knowledge (Sec. 2.1), we replace these with different projection operations via asymmetric convolutions. More specifically, we parametrize ***U*** and ***V*** as convolutional layers with kernel size (1, *kernel size), (kernel size*, 1) and stride of (1, 2), (2,1), respectively. As a result, 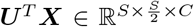, and focuses on the contextual information along each vertical column. 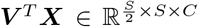, and focuses on the contextual information of the horizontally layered structures. Apart from these two structure-orientated asymmetrical convolutional operations, we also replace the original linear projection ***P*** with a third parametrized convolutional operation for feature extraction. This operation is adopted as two consecutive standard convolutional blocks; each convolutional block consists of a convolutional layer followed by a Batch Normalisation [20] and a ReLU [21]. This operation generates features as 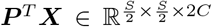. The two streams of the bilinear model are multiplied together using the Hadamard product after a transpose operation to match their shapes. We, then, reshape the feature to 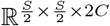 to match the shape of tensor ***P****^T^X*. Finally, we apply a Sigmoid function for normalization to generate an attention mask, which is then combined to the result of the third feature extraction stream. The higher-order low-rank attention is then given by:

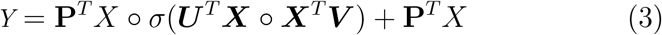

To further increase modelling efficiency and capacity, we apply a multi-scale strategy and multi-grouped channels: Let {*P_i_*} _i=i,…,4_ be sets of asymmetrical convolutional layers with kernel size of (1, 2), (1, 3), (1, 5), (1, 7), respectively and *C_out_* the output channel number. Then, ∀ {*P_i_*}*_i_*_=1,…,4_ ∃ different numbers of groups of filters at *C_out_//*8, *C_out_//*4, *C_out_//*2, *C_out_*. Our proposed attention model is finally:

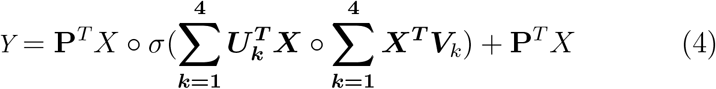

### 2.3 Architectural Overview

Our architecture consists of three branches: an encoder-decoder main branch and two parallel attention side branches. Hereinafter, the main branch is referred to as backbone. The backbone captures multi-scale visual features and integrates low-level features with high-level ones, whereas the two side branches attend to the horizontal and vertical directions. The two side branches calculate the attention weights as described in Sec. 2.2. Fig. 1 illustrates the proposed framework. Our proposed architecture is composed of downsampling and upsampling components, each alternating between a convolutional block and an oriented attention block. Each downsampling block halves the size of the feature maps in height and width and doubles the channels, while each upsampling block doubles the feature maps in height and width while halving the channels.

**Fig. 1:**
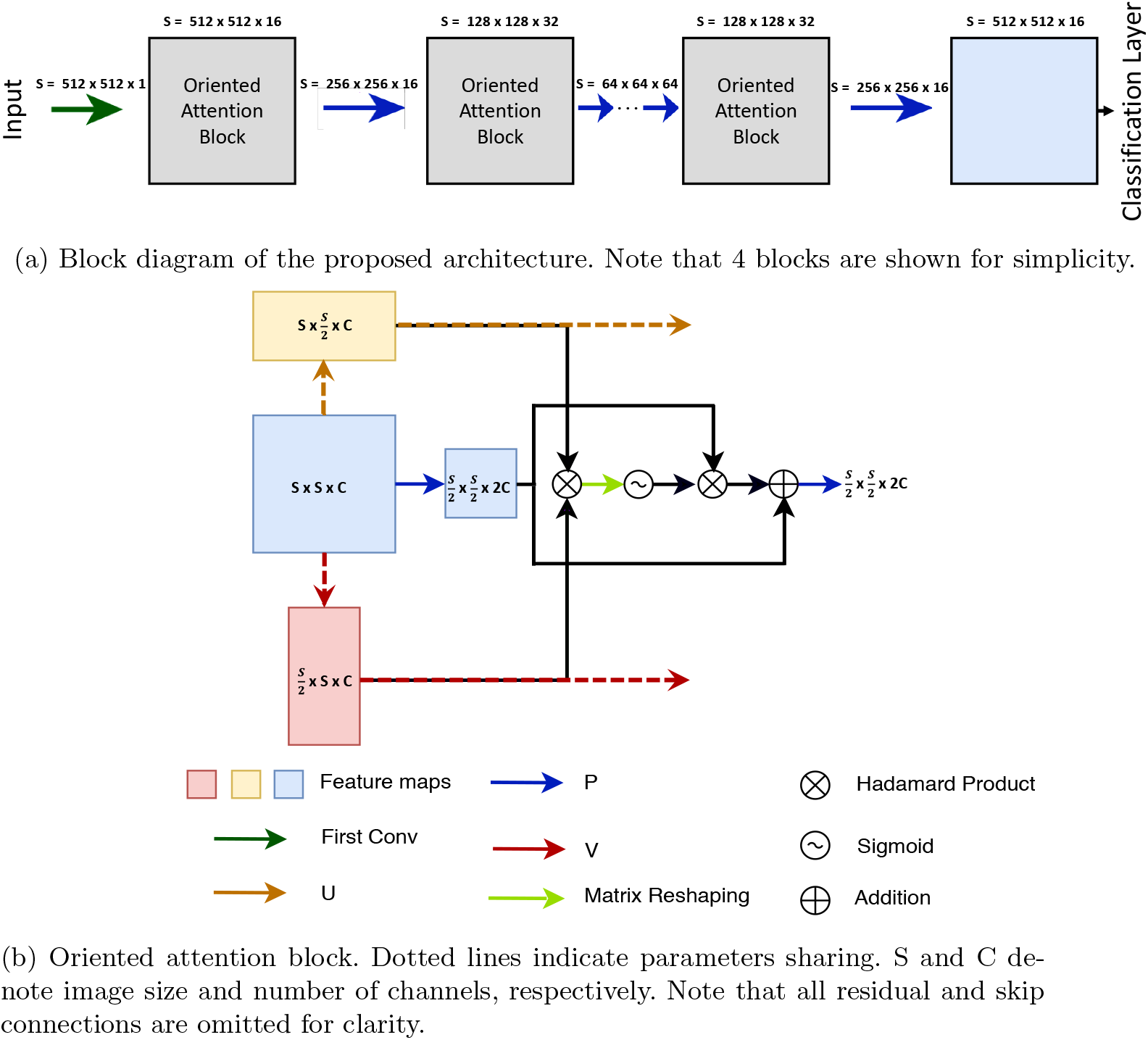
Illustration of the proposed methodology.

## 3 Experiments and Results

### 3.1 Data

We used two clinical studies, COMPASS [15] and RAPID [1], and the publicly available Duke dataset [3] to evaluate our proposed methodology, conducting both binary and multi-class segmentation. All acquisitions are circular OCT (496 × 796) scans. Note that our method’s ability to segment all retinal layers is illustrated in the Duke dataset; eight boundaries are annotated. The precision, repeatability and agreement of our method are evaluated independently on COMPASS and RAPID, using one layer, i.e. RNFL.

A. Block diagram of the proposed architecture. Note that 4 blocks are shown for simplicity.
B. Oriented attention block. Dotted lines indicate parameters sharing. S and C denote image size and number of channels, respectively. Note that all residual and skip connections are omitted for clarity.

**RAPID study** The RAPID study consists of 82 stable glaucoma patients attended Moorfields Eye Hospital for up to 10 visits within a 3-month period, consisting of 502 SDOCT (SpectralisOCT, Heidelberg Engineering) images. We split the RAPID study into training, validation and testing images [1].

**COMPASS study** To test the generalizability of our method, we evaluate the trained models from RAPID on unseen cases from COMPASS. The COMPASS study consists of 943 subjects (499 patients with glaucoma and 444 healthy subjects), attended multiple centres for up to 2 years consisting of 931 SDOCT (SpectralisOCT, Heidelberg Engineering) images [15].

**Duke dataset** The Duke dataset [3] consists of 110 annotated SDOCT obtained from 10 patients suffering from Diabetic Macular Edema (DME) (11 B-scans per patient). Scans were annotated by two experts. They include: region above the retina (RaR), inner limiting membrane (ILM), nerve fiber ending to inner plexiform layer(NFL-IPL), inner nuclear layer (INL), outer plexiform layer (OPL), outer nuclear layer to inner segment myeloid (ONL-ISM), inner segment ellipsoid (ISE), outer segment to retinal pigment epithelium (OS-RPE) and region below the retina (RbR). Note that segmenting fluid is beyond the scope of this work.

### 3.2 Experimental Setup

To illustrate the effectiveness of our model-agnostic LROA modules, we compare each LROA-enhanced network with the corresponding backbone architecture. We use the following models that have been shown to perform well on retinal OCT segmentation tasks: U-Net [14], SegNet [22], DRUNET [13] and ReLayNet [11] to prove our hypothesis. Since LROA is based on attention mechanisms, we further include a state-of-the-art attention enhanced network, namely Attention-Unet [23]. All baselines models are re-implemented in an identical fashion as the respective papers, without pre-training, for fair comparison. Henceforth, the network using U-net [14] as backbone is referred to as “LROA-U”, the network using SegNet [22] as backbone is referred to as “LROA-S”, the network using DRUNET [13] as backbone is referred to as “LROA-D” and the one using RelayNet [11] as backbone is referred to as “LROA-R”. We also implement two versions of LROA-S with different sized kernels in {*P_i_*}*_n_*_=1,…,4_ and {*V_i_*}*_n_*_=1,…,4_ to investigate the effect of size kernel. The first variant of LROA-S uses a larger kernel with a size of (1, 3), (1, 5), (1, 7) and (1, 9) in {*P_i_*}*_n_*_=1,..,4_, and is referred to as “LROA-SL”. The second variant of LROA-S uses a larger kernel size of (1, 3), (1, 7), (1, 9) and (1,15) in {*P_i_*}*_n_*_=1,…,4_, and is referred to as “LROA-SVL”. To quantify the relative diagnostic precision, repeatability and test-retest variability, we test, independently, one layer (RNFL) as done in similar studies, i.e. predicted versus ground truth RNFL thickness on RAPID and COMPASS. To illustrate the method’s segmentation improvement, we segment all layers on the Duke dataset, but the fluid region, which is beyond the scope of this work due to the very limited number of training images. All experiments are patient-independent.

**Training** All images were resized to 512 × 512. Training images are augmented with random probability using channel ratio modification, horizontal and vertical flipping and Gaussian and speckle noise corruption. We use Standard crossentropy loss, AdamW optimizer [24], an initial learning rate of 10^-3^, and a minibatch size of 4 until convergence, across all experiments. All experiments were performed on a NVIDIA Titan V (12GB) GPU using PyTorch. Code is publicly available at github.com/gelazari/MICCAI2020.

### 3.3 Results

Tables 1, 2 and Fig. 2 illustrate our results. Our approach improves across all experiments. Table 1 shows the 95% limits of agreement (LOA), mean difference, and the mean standard deviation (SD) of the difference for three visits across all subjects on the RAPID and COMPASS study. Following similar studies, we use the average RNFL segmentation to compute these metrics. The results show that our approach outperforms all other methods: diagnostic precision and repeatability are markedly improved. Moreover, our method not only produces segmentations with high ground truth agreement, but also reduces test-retest variability. Importantly, we appreciate a statistically significant improvement in the aforementioned metrics obtained with LROA-U (best of proposed submodels) as compared to those obtained with U-Net (best baseline)(p = 0.037, Mann-Whitney U test), leading to a lower sample size in a clinical trial power analysis. Fig. 2 illustrates the corresponding Bland-Altman plots; LROA leads to significantly better agreement and lower test-retest variability. Table 2 shows multi-class segmentation results on the Duke dataset, including the positive impact from larger sized asymmetrical kernels. It can be seen that the proposed method outperforms all the others by huge margins. For instance, LROA-S improves over its backbone SegNet by 55% in IoU. Fig. 3 shows visual segmentation results. Note that segmenting fluid is beyond the scope of this work.

**Fig. 2:**
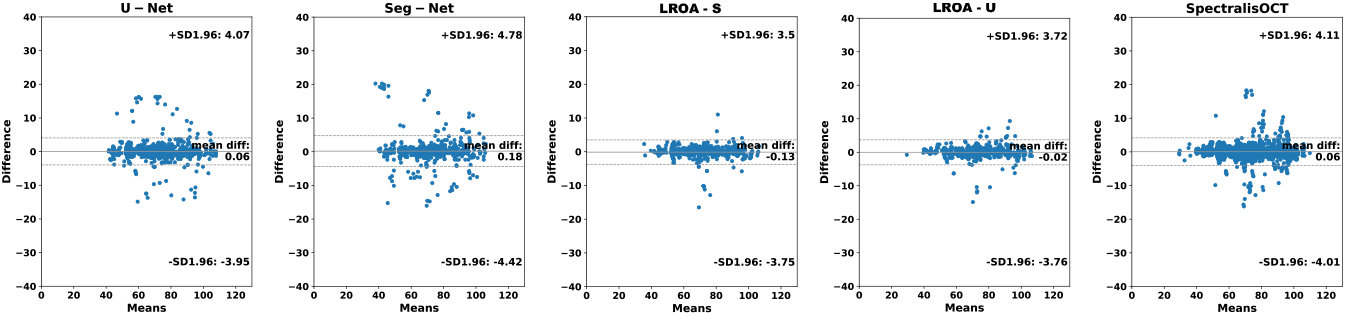
Bland-Altman plots between all methods and ground truth. Note that for SpectralisOCT, repeated test-retest measurements for each eye are used.

**Fig. 3:**
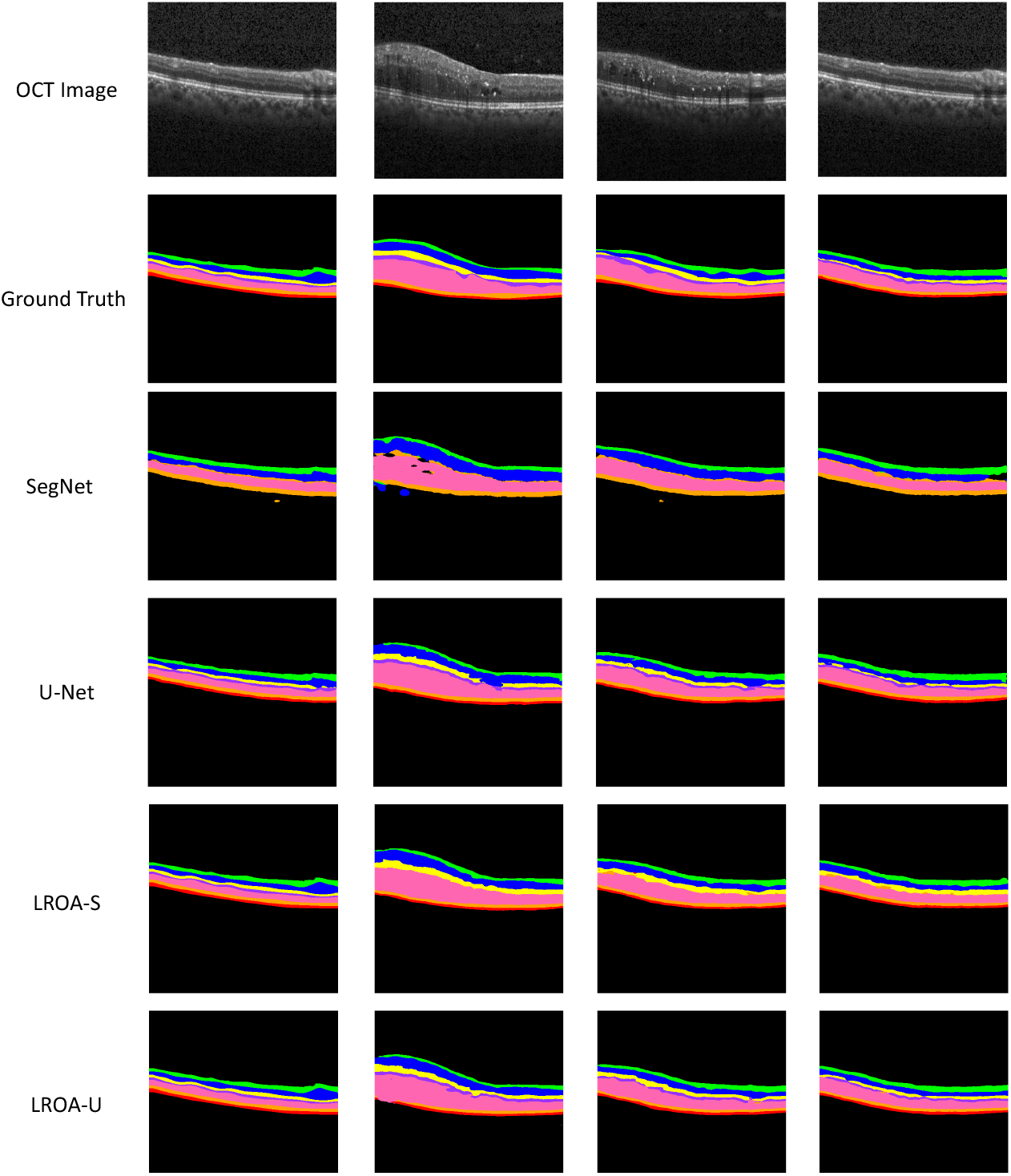
Segmentation results on the Duke dataset.

**Table 1:** Limits of agreement, mean difference of all methods versus ground truth, and mean SD (test-retest variability) of the first three visits difference. Results on binary RNFL segmentation on COMPASS and RAPID studies.

| Method | SegNet | **LROA-S** | U-Net | **LROA-U** | SpectralisOCT |
| --- | --- | --- | --- | --- | --- |
| 95% LOA | [4.78, -4.42] | **[3.50, -3.75]** | [4.07, -3.95] | [3.72, -3.76] | [4.70, -4.48] |
| Mean Diff. | 0.18 | -0.13 | 0.06 | **-0.02** | 0.11 |
| Mean SD | 1.93 | 1.22 | 1.84 | **1.13** | 1.20 |

**Table 2:** Multi-Class segmentation results on the Duke dataset.

| Networks | IoU (%) | F1 (%) | Recall (%) | Precision (%) | MSE |
| --- | --- | --- | --- | --- | --- |
| SegNet | 45.22 | 52.98 | 58.68 | 53.17 | 0.94 |
| **LROA-S** | 70.20 | 82.33 | 84.43 | 81.98 | 0.31 |
| **LROA-SL** | 75.99 | 88.40 | 89.97 | 88.63 | 0.28 |
| **LROA-SVL** | 76.68 | 89.08 | 90.42 | **92.81** | **0.26** |
| U-Net | 69.08 | 81.47 | 83.38 | 80.77 | 0.39 |
| Atten-UNet | 70.52 | 83.52 | 85.07 | 83.16 | 0.41 |
| **LROA-U** | 76.65 | 89.07 | 90.03 | 89.48 | 0.30 |
| ReLayNet | 74.80 | 87.64 | 88.49 | 87.99 | 0.36 |
| **LROA-R** | **78.59** | **91.27** | **91.57** | 92.25 | 0.29 |
| DRUNET | 69.73 | 81.97 | 83.81 | 81.12 | 0.39 |
| **LROA-D** | 75.77 | 88.22 | 89.79 | 89.31 | 0.31 |

## 4 Discussion and Conclusion

In this paper, we present a novel, end-to-end trainable, attentive model for retinal OCT segmentation. Our contributions extend current literature as we highlight valuable features of high-level layers, efficiently combined with high-order attention information in two relevant dimensions, to guide the final segmentation. Our approach is based on feature correlation learning, exploiting the horizontally-layered retinal structure and the vertical partitioning of retinal surfaces. The proposed methodology appears robust and flexible in terms of capacity and modularity. Results show the model not only significantly improves segmentation results, but can also increase the statistical power of clinical trials with layer thickness change outcomes. Future work will focus on integrating context among different B-scans.

## Data Availability

All data used are publically available data.

